# Case fatality rate in COVID-19: a systematic review and meta-analysis

**DOI:** 10.1101/2020.04.01.20050476

**Authors:** Chanaka Kahathuduwa, Chathurika Dhanasekara, Shao-Hua Chin

## Abstract

**Background:** Estimating the prevalence of severe or critical illness and case fatality of COVID-19 outbreak in December, 2019 remains a challenge due to biases associated with surveillance, data synthesis and reporting. We aimed to address this limitation in a systematic review and meta-analysis and to examine the clinical, biochemical and radiological risk factors in a meta-regression.

**Methods:** PRISMA guidelines were followed. PubMed, Scopus and Web of Science were searched using pre-specified keywords on March 07, 2020. Peer-reviewed empirical studies examining rates of severe illness, critical illness and case fatality among COVID-19 patients were examined. Numerators and denominators to compute the prevalence rates and risk factors were extracted. Random-effects meta-analyses were performed. Results were corrected for publication bias. Meta-regression analyses examined the moderator effects of potential risk factors.

**Results:** The meta-analysis included 29 studies representing 2,090 individuals. Pooled rates of severe illness, critical illness and case fatality among COVID-19 patients were 15%, 5% and 0.8% respectively. Adjusting for potential underreporting and publication bias, increased these estimates to 26%, 16% and 7.4% respectively. Increasing age and elevated LDH consistently predicted severe / critical disease and case fatality. Hypertension; fever and dyspnea at presentation; and elevated CRP predicted increased severity.

**Conclusions:** Risk factors that emerged in our analyses predicting severity and case fatality should inform clinicians to define endophenotypes possessing a greater risk. Estimated case fatality rate of 7.4% after correcting for publication bias underscores the importance of strict adherence to preventive measures, case detection, surveillance and reporting.

## Introduction

A novel corona virus, first identified in Wuhan, China in late 2019, resulted in a pandemic by the first quarter of 2020, contributed by the prolonged survival of the virus in the environment and extended length of pre-or post-symptomatic and potential asymptomatic shedding. ^1-4^ While the virus is known to cause only a mild illness in a majority, severe illness characterized by respiratory distress requiring hospital admission is not uncommon. ^5^ Furthermore, the virus has the potential to precipitate a life-threatening critical illness, characterized by respiratory failure, circulatory shock, sepsis or other organ failure, requiring intensive care. ^6, 7^

An extensive body of literature that emerged since the outset of the epidemic in China, have examined the rates of severe and critically severe illness as well as case fatality associated with COVID-19. However, the literature on COVID-19 has several limitations. First, due to lack of awareness and limited availability of training and resources to confirm the diagnosis, failure to recognize and code COVID-19 as the potential cause of morbidity and mortality may have contribute to under estimation of the effects of COVID-19. ^8^ In fact, a recent estimate suggested that approximately 86% cases of COVID-19 were not documented prior to January 23, 2020. ^4^ On the contrary, screening of only those who are at high risk may lead to over-reporting of morbidity and mortality. Second, given that most datasets and publications are derived from retrospective chart review, as opposed to prospective methods, high measurement error is inevitable. ^9^ Third, the literature on the outcomes are originating mainly from tertiary care settings, distorting the overall clinical picture. ^10^ Finally, including the same patients in multiple reports examining the same research question without clearly indicating is a major lapse in methodological and ethical standards. ^11^

As such, we aimed to conduct a systematic review of the available literature to identify publications with minimal potential overlap, in order to estimate the prevalence of severe illness, critical illness and case fatality among individuals with COVID-19 in random-effects meta-analyses to enhance generalizability. More importantly, we aimed to adjust our prevalence estimates by correcting for publication bias and underreporting. We further aimed to examine the effects of clinical, biochemical and radiological risk factors moderating the between-study heterogeneity of the severity and case fatality rates.

## Methods

### Search strategy and selection criteria

All procedures were conducted in accord with the Preferred Reporting Items for Systematic Reviews and Meta-analyses (PRISMA) guidelines. PubMed, Scopus and Web of Science databases were searched on March 7, 2020 with the aim of identifying studies that have been published in year 2020 examining the prevalence of severe illness, critically severe illness and mortality associated with COVID-19 infection using pre-determined keyword combinations (Table S1 in Appendix). No language restrictions were applied. Duplicate records were removed and titles and abstracts were screened for pre-defined eligibility criteria (Figure 1) by two independent raters (CD and CK or SC). Records published in Chinese were translated to American English using Google translator and a native Chinese speaker (SC) examined the original records in Chinese. Full-text manuscripts of records that were deemed eligible in the initial screening were re-examined. All eligible full-text manuscripts were examined using the 9-item Quality Assessment Tool for Case Series Studies of the National Heart, Lung and Blood Institute by two study personnel.

**Figure 1:**
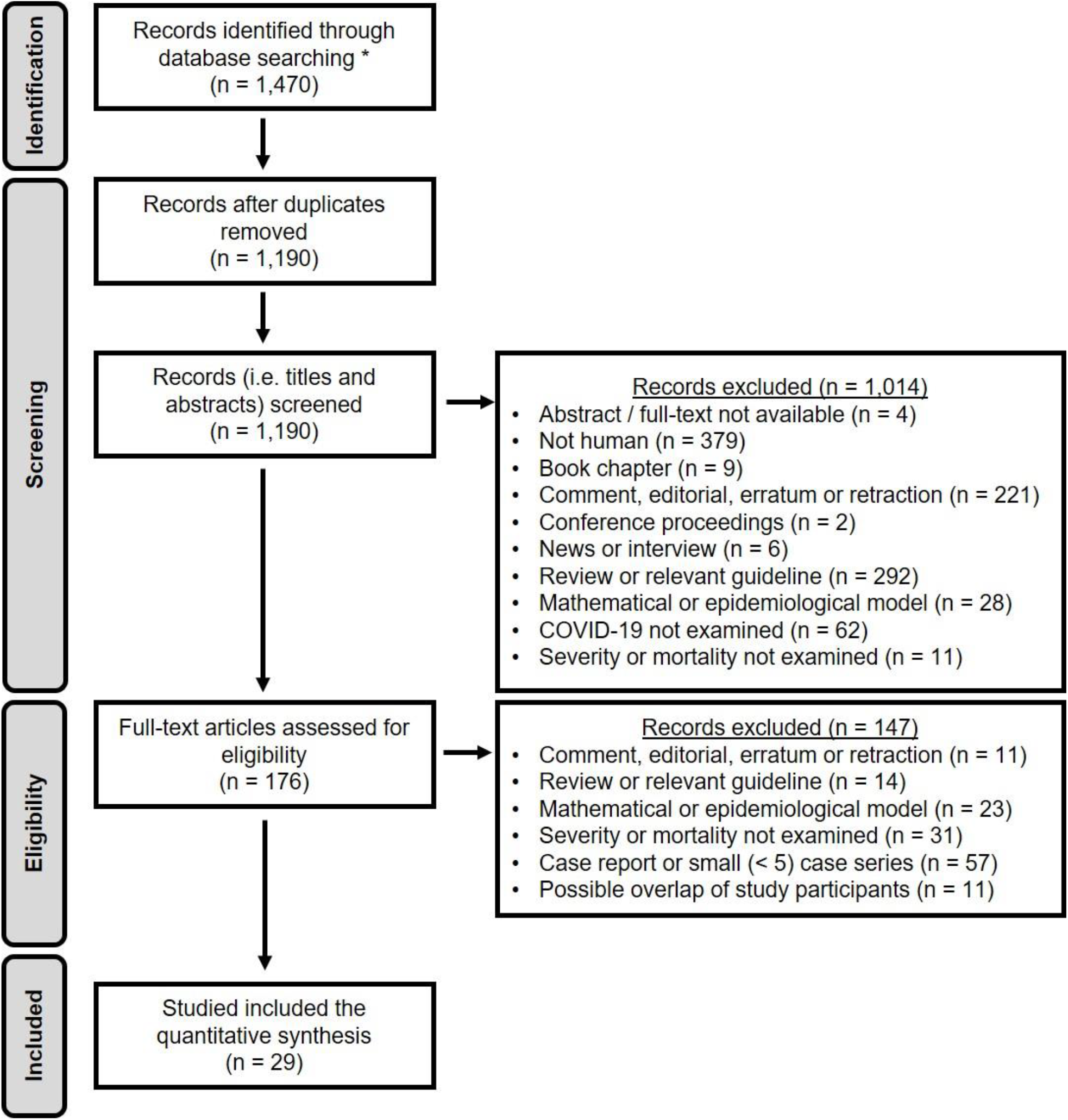
PRISMA flow chart outlining eligibility criteria and study selection.

Data were extracted from the eligible manuscripts into pre-defined data-fields. The data fields included the total sample size and number of participants with severe illness, critically severe illness and mortality. Severe illness was operationally defined as having either respiratory distress with RR ≥ 30 /min, resting peripheral oxygen saturation of ≤ 93% or arterial partial pressure of oxygen ≤ 300 mmHg, requiring hospitalization. Critical illness was defined as having respiratory failure, circulatory shock, end-organ failure or any combination of the above requiring intensive care. In addition, the following variables were extracted as potential covariates of the above outcomes. Central tendency (i.e. mean or median) and dispersion (i.e. SD, SE, 95%CI, IQR or range) of age were extracted. When not reported, study level means and standard deviations for age were imputed from the available statistics (i.e. median, IQR or range). ^12^ Proportions of the following variables within a study sample were extracted: age ≤ 18 years, age ≥60 years, female sex, diabetes mellitus, hypertension, heart disease, chronic liver disease, chronic kidney disease, chronic obstructive pulmonary disease, malignancy, immunosuppression (e.g. HIV), smoking and pregnancy. Proportions of patients with specific presenting symptoms (i.e. fever, cough, sore throat, shortness of breath, headache, diarrhea), asymptomatic cases, specific laboratory parameters (i.e. positive nucleic acid test for COVID-19, leukopenia, leukocytosis, thrombocytopenia, lymphopenia, elevated lactate dehydrogenase (LDH), elevated C-reactive protein (CRP), elevated erythrocyte sedimentation rate (ESR), high procalcitonin and high D-dimer based on reference ranges considered in each study) and radiographic features (i.e. no lesions on CT, patchy consolidation, ground glass opacities, peripheral distribution, and bilateral lung involvement or involvement of ≥ 3 lobes).

### Data analysis

Three separate DerSimonian-Laird random-effects meta-analyses were performed using the ‘meta’ package (version 4.11-0) in R statistical software (version 3.6.2) to examine three primary outcomes: the prevalence of a) combined severe or critical COVID-19 infection, b) critically severe COVID-19 infection, and c) COID-19-associated mortality. ^13^ Studies with both zero or 100% proportions were not excluded to ensure incorporation of all available data, which is also known to ensure analytic consistency and minimize bias. ^14^ Consistency of the findings of the meta-analyses were confirmed by leave-one-out sensitivity analyses. ^15^ Given that under-reporting and publication bias could result in biased (i.e. smaller) prevalence estimates, publication bias was examined using funnel plots and the effect-sizes were imputed for estimated missing (i.e. unpublished / unreported) studies via the trim-and-fill method. ^16, 17^ The meta-analyses were repeated including the effect-size estimates of these potentially missing studies in order to obtain unbiased estimates of the three primary outcomes. Heterogeneity of effect-sizes was quantified by calculating the Higgins’ I^2^ statistic for each meta-analysis. ^18, 19^ To explain the heterogeneity of the studies ^20^, exploratory univariate random-effects meta-regression analyses were performed to examine the moderator effects of each of the covariates described above.

## Results

Results of database search, subsequent screening and eligibility assessment are summarized in a PRISMA flow diagram (Figure 1). Out of 1,470 records identified in the initial database search, 29 studies including data of 2,090 patients with COVID-19 were deemed eligible. The proportions of females in the study samples ranged from 27.59-100.00%. The mean age of the participants included in the studies ranged from 2-66 years. Four studies recruited entirely on children and one study exclusively recruited pregnant women. The included studies and their quality ratings are summarized in Table S2 with their references. The summary statistics of all covariates are summarized in Table S3 in Appendix.

Pooled prevalence of severe or critically severe illness among individuals with COVID-19 infection was estimated to be 14.6% (95%CI, 8.9%-23.1%) in the random-effects meta-analysis (Figure 2a). Excluding any single study from the meta-analysis (i.e. leave-one-out sensitivity analyses) did not significantly change the pooled prevalence estimate. However, funnel plot of the effect-sizes was severely asymmetric, suggesting substantial underreporting or publication bias (Figure 2b). Seven effect-sizes were imputed to correct for the publication bias. When the random-effects meta-analysis was performed including these imputed effect-sizes (i.e. after correcting for publication bias), the prevalence of severe or critical illness increased to 25.8% (95%CI, 17.2%-36.8%) (Figure 2c).

**Figure 2:**
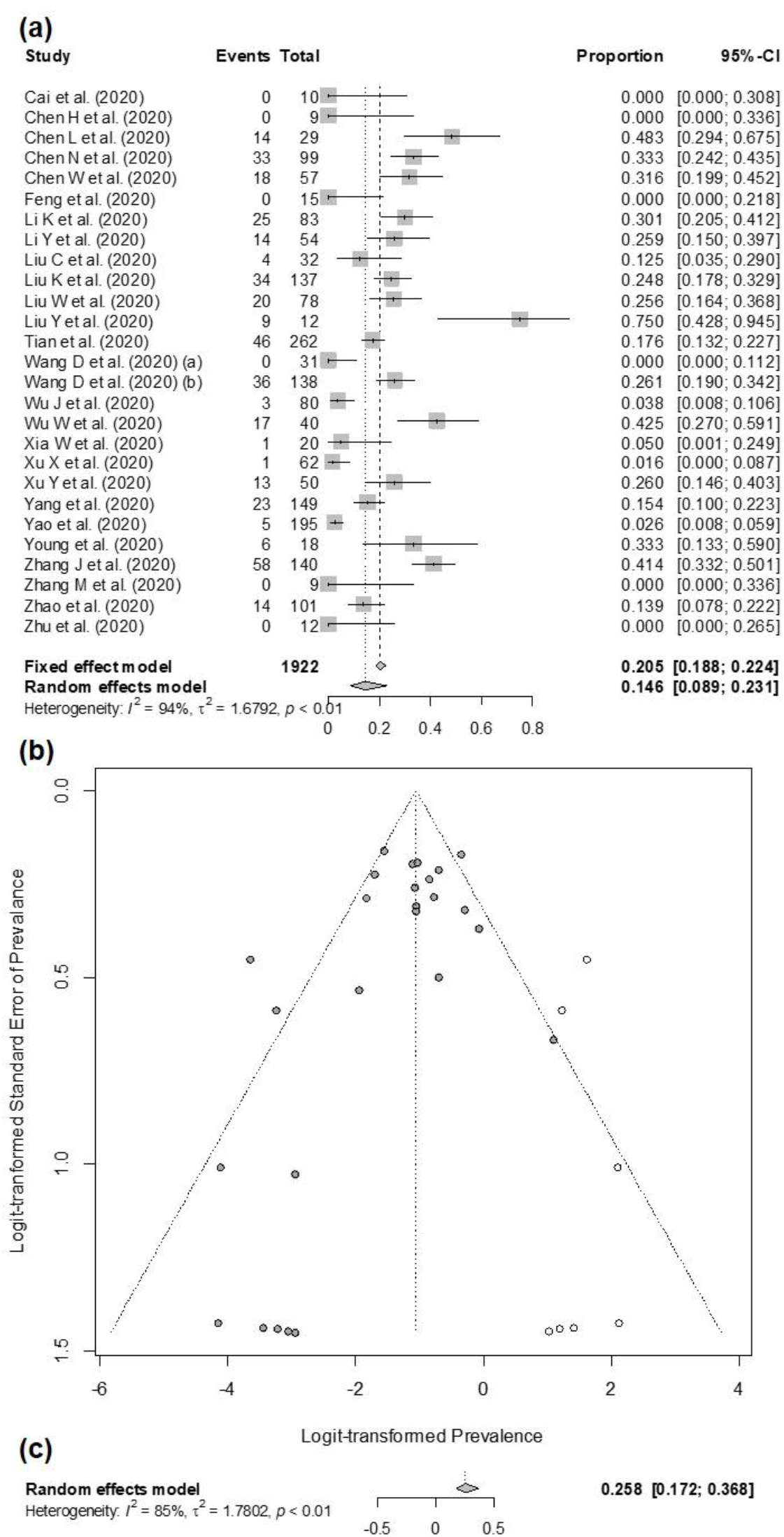
Results of random-effects meta-analysis examining the pooled prevalence of combined severe and critical illness among individuals with COVID-19. a) Forest plot; b) Funnel plot depicting publication bias and imputed effect-sizes to correct for publication bias; c) Results corrected for publication bias.

Significant heterogeneity was observed among the prevalence estimates of severe illness (τ^2^ = 1.679; I^2^ = 94%, p < 0.001). Correcting for publication bias decreased this heterogeneity (I^2^ = 85%, 95%CI, 80%-89%), however, heterogeneity remained significant (p < 0.001). Exploratory univariate random-effects meta-regression analyses conducted with the aim of explaining the heterogeneity using the moderator effects of the considered covariates suggested that each of increasing mean age (p = 0.006) and prevalence of age ≥ 60 years (p < 0.001), hypertension (p < 0.001), chronic kidney disease (p = 0.038), malignancy (p = 0.023) and chronic obstructive pulmonary disease (p = 0.025) were associated with a greater risk of severe or critical illness associated with COVID-19, while the prevalence of age ≤ 18 years (p = 0.007) within a sample was associated with a reduced risk of severe or critical illness. Prevalence of the presenting clinical features of fever (p < 0.001), dyspnea (p = 0.028) and diarrhea (p = 0.026); laboratory findings of lymphocytopenia (p = 0.003), elevated LDH (p < 0.001), CRP (p < 0.001) and D-dimer levels (p < 0.001); and bilateral lung involvement or involvement of ≥ 3 lung lobes (p = 0.006) was associated with increased risk of severe or critically severe illness, while having no radiological features on chest CT was associated with decreased risk of severe illness (p = 0.003). The results of all univariate meta-regression analyses examining the moderator effects of the covariates on the prevalence of severe or critical illness in COVID-19 infection and their effects on heterogeneity are summarized in Table S4 in Appendix.

The random effects meta-regression analyses revealed a pooled estimate of 4.8% (95%CI, 2.4%-9.5%) for the prevalence of critical illness in COVID-19 infection (Figure 3a). Leave-one-out meta-regression analyses did not significantly change this estimate. The funnel plot of effect-sizes (Figure 3b) was highly suggestive of underreporting or publication bias. Eleven effect-sizes had to be imputed to statistically correct for this bias and after correction, the pooled prevalence of critical illness in COVID-19 infection increased to 16.3% (95%CI, 9.8%-25.7%) (Figure 3c).

**Figure 3:**
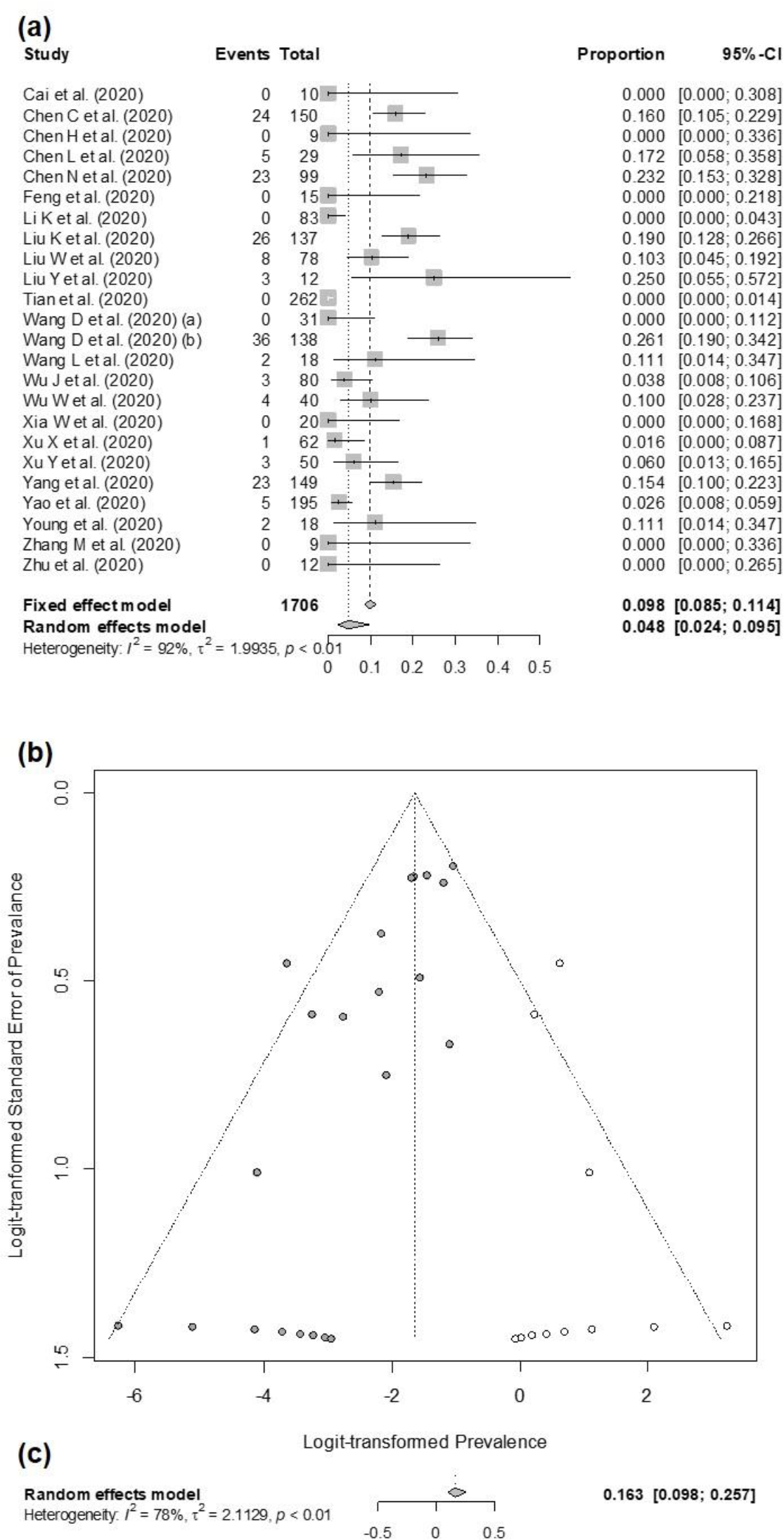
Results of random-effects meta-analysis examining the pooled prevalence of critical illness among individuals with COVID-19. a) Forest plot; b) Funnel plot depicting publication bias and imputed effect-sizes to correct for publication bias; c) Results corrected for publication bias.

Significant heterogeneity of effect-sizes was a concern for the meta-analysis of prevalence of critical illness as well (τ^2^ = 1.994; I^2^ = 92%, p < 0.001). Correcting for publication bias decreased this heterogeneity (I^2^ = 78%, 95%CI, 69%-84%), however, heterogeneity remained significant (p < 0.001). Univariate meta-regression analyses suggested increased risk of critical illness associated with sample characteristics of increasing mean age (p = 0.002), prevalence of age ≥ 60 years (p < 0.001), comorbid hypertension (p < 0.01), cardiac disease (p = 0.023) and malignancy (p = 0.041). Similarly, prevalence of fever (p = 0.044), dyspnea (p = 0.042) and fatigue (p = 0.036) on presentation; prevalence of increased LDH (p = 0.003), CRP (p = 0.008) and D-dimer (p = 0.021) were associated with a greater risk of critical illness (Table S5 in Appendix).

Prevalence of COVID-19 associated mortality was 0.8% (95%CI, 0.2%-2.9%) based on the random-effects meta-analysis (Figure 4a) and this estimate was minimally affected by leave-one-out sensitivity analyses. However, as with severe and critically severe illness, publication bias / potential underreporting was apparent on the funnel plot of effect sizes (Figure 4b). Thirteen effect-sizes were imputed to account for missing / unreported effects in an attempt to statistically correct for the publication bias and conducting the meta-regression analyses with the addition of these effect-sizes revealed a pooled estimate of 7.4% (95%CI, 4.5%-11.9%) for the mortality rate associated with COVID-19 infection (Figure 4c).

**Figure 4:**
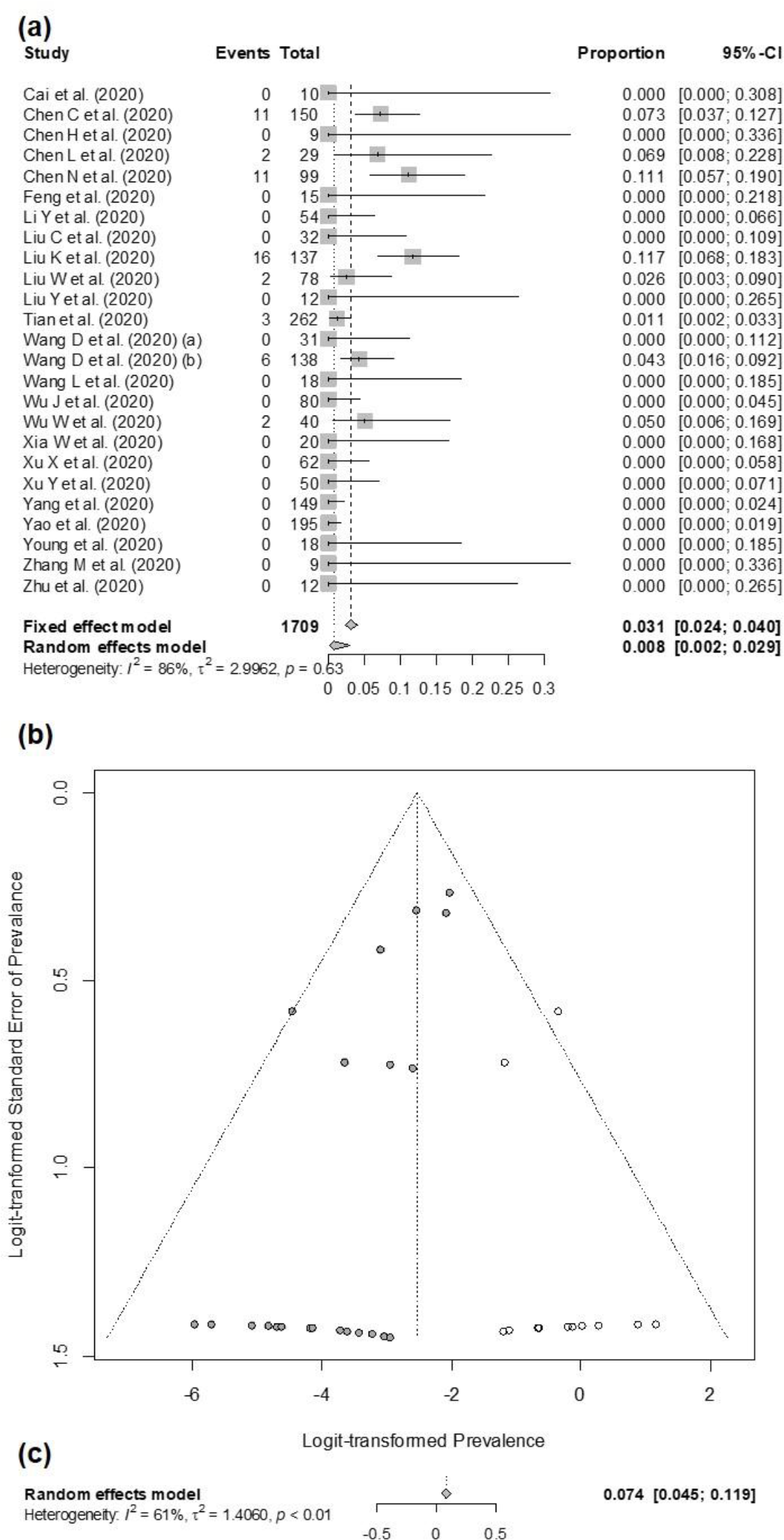
Results of random-effects meta-analysis examining the pooled case fatality rate among individuals with COVID-19. a) Forest plot; b) Funnel plot depicting publication bias and imputed effect-sizes to correct for publication bias; c) Results corrected for publication bias. DerSimonian R, Laird N. Meta-analysis in clinical tria

Heterogeneity of effect-sizes on prevalence of mortality in COVID-19 was also a concern (τ^2^ = 2.996; I^2^ = 86%, p < 0.001). Correction for publication bias decreased the heterogeneity (I^2^ = 61%, 95%CI, 45%-73%), yet the heterogeneity remained significant (p < 0.001). Univariate meta-regression analyses modeling heterogeneity indicated increased mortality risks associated with increasing mean age (p < 0.001), prevalence of age ≥ 60 years (p = 0.011), presenting with fatigue (p = 0.048), leukocytosis (p =0.007), high LDH (p = 0.030) and low albumin (p < 0.001). Prevalence of age ≤ 18 years (p = 0.036) was associated with a decreased risk of COVID-19-associated mortality (Table S6 in Appendix).

## Discussion

In this systematic review and meta-analysis, we comprehensively and systematically examined the available literature to estimate the prevalence of morbidity and mortality associated with SRAS-CoV-2 infection. Despite the large number of articles that were reviewed, our quantitative synthesis was limited to 29 studies representing data of 2,090 individuals. Our literature-based estimates of severe illness, critical illness and case fatality rates among patients with COVID-19 were 15%, 5% and 0.8% respectively. After adjusting for underreporting and publication bias, COVID-19-associated prevalence of severe illness, critical illness and case fatality increased to 26%, 16% and 7.4% respectively.

Our unadjusted random-effects estimates of severe illness requiring hospitalization (15%) and critical illness requiring intensive care admission (5%), are consistent with the estimates of COVID-19-associated morbidity based on large individual-level datasets. ^21^As such, the unadjusted findings of our meta-analysis regarding severity of illness corroborate the inferences made based on current surveillance systems. However, the unadjusted mortality rate observed in our analysis (0.8%, 95% CI, 0.2%-2.9%) is lower than the COVID-19-associated mortality rates in China (3.6%) or globally (3.4%) at the end of February, 2020 (i.e. the time represented in the reviewed publications). ^22^

As we have noted in the introduction, retrospective patient data and the literature derived from such data could be systematically biased towards both overestimating and / or underestimating morbidity and mortality. The reviewed studies are largely representing tertiary care settings, of which the capacity may have been overridden minimally, if at all, despite the high reproductive number (R_0_) at the time of sampling. As such, the outcomes of our unadjusted random-effects meta-analyses, which accounts for the random variability of effects between studies, can be inferred to be a generalizable representation of morbidity and mortality rates applicable to a well-trained and equipped healthcare setting of which resources are not overwhelmed. This estimate therefore represents COVID-19 associated mortality in regions with a low R_0_.

However, reverse-causation bias caused by failing to capture the deaths that may not reach healthcare facilities / be diagnosed prior to death may contribute to underestimation. ^23^ The effect-sizes imputed to correct for publication bias may in fact represent severity and case fatality rates of settings where the demand exceeds the available resources. ^24^ In fact, our data substantiate the recent forecasts and recommendations to suppress the epidemic growth. ^25^ While optimized utilization of healthcare facilities by maintaining a low R_0_ may reduce the mortality rates to as low as 0.8%, overwhelming healthcare resources may increase the overall case fatality rate to 7.4%, or even greater as represented by the effect-sizes we have imputed. ^26^

In our meta-regression analyses that examined risk factors, increasing age and age ≥ 60 years consistently stood out as a risk factor, while age ≤ 18 years consistently remained a protective factor, being consistent with the current literature. Angiotensin converting enzyme (ACE)-2 receptors are generally upregulated in patients with hypertension or heart failure, who receive ACE-inhibitors and angiotensin-II receptor blockers. ^27^ COVID-19 is known to enter cells by binding to ACE-2 receptors, increasing their risk of infection and development of severe clinical illness. ^27^ Consistently, we observed an increased risk of severity with hypertension and cardiovascular disease. Presenting with fever emerged as a risk factor for severe and critical illness as well as mortality, underscoring the importance body temperature as a screening tool. ^28^

Our systematic review has some limitations. First, while we eliminated studies with overlapping samples by screening for overlaps of institutes, study dates and authors, we cannot be 100% certain. Second, high degree of heterogeneity was a concern. However, this should be expected in any meta-analysis due to the variability in methodology and study samples and we used heterogeneity to explore covariates in meta-regression analyses. ^18, 20^ Third, funnel plots of all three meta-analyses indicating substantial publication bias, limiting the generalizability of uncorrected random-effects meta-analyses. Finally, the protocol was not pre-registered. Use of a systematic search strategy; use of random-effects meta-analyses and meta-regression analyses assuming high heterogeneity of effect-sizes; exploring the etiology of heterogeneity in meta-regression analyses, which also identified risk factors of morbidity and mortality; exploring the validity of our findings in sensitivity analyses; and statistically correcting for publication / underreporting bias are notable strengths of our systematic review and meta-analysis.

In conclusion, after correcting for publication bias, COVID-19 associated overall rates of requirement for hospitalization, intensive care and case fatality could be as high as 26%, 16% and 7.4% respectively. This underscores the importance of strict adherence to preventive measures, case detection, surveillance and reporting. Hypertension; fever and dyspnea at presentation; and elevated CRP seem to predict increased disease severity, while increasing age and elevated LDH seem to consistently predict severity and case fatality. These risk factors should inform clinicians to define endophenotypes possessing a greater risk.

## Data Availability

The data presented in the manuscript is based entirely on data extracted from peer-reviewed, published research. The authors are willing to share detailed records of all steps including searching databases, systematically screening the literature, quality assessment, data extraction and meta-analysis (including the scripts).

## Contributors

CK, CD and SC conceptualized the study; CK and CD conducted the literature search; CK, CD and SC were involved in screening the literature; CK and CD extracted the data; CK performed the meta-analyses and meta-regression analyses; CK and CD wrote the manuscript; all authors read and agreed to the content of the manuscript.

## Supplementary Appendix

**Table S1:** Keywords and keyword combinations used to screen the PubMed, Scopus, and Web of Science electronic databases

|  |
| --- |
| (covid-19) OR (((corona OR coronavirus)) AND wuhan) OR (2019 novel coronavirus infection) OR (COVID19) OR (coronavirus disease 2019) OR (coronavirus disease-19) OR (2019-nCoV disease) OR (2019 novel coronavirus disease) OR (2019-nCoV infection) |

**Table S2:**
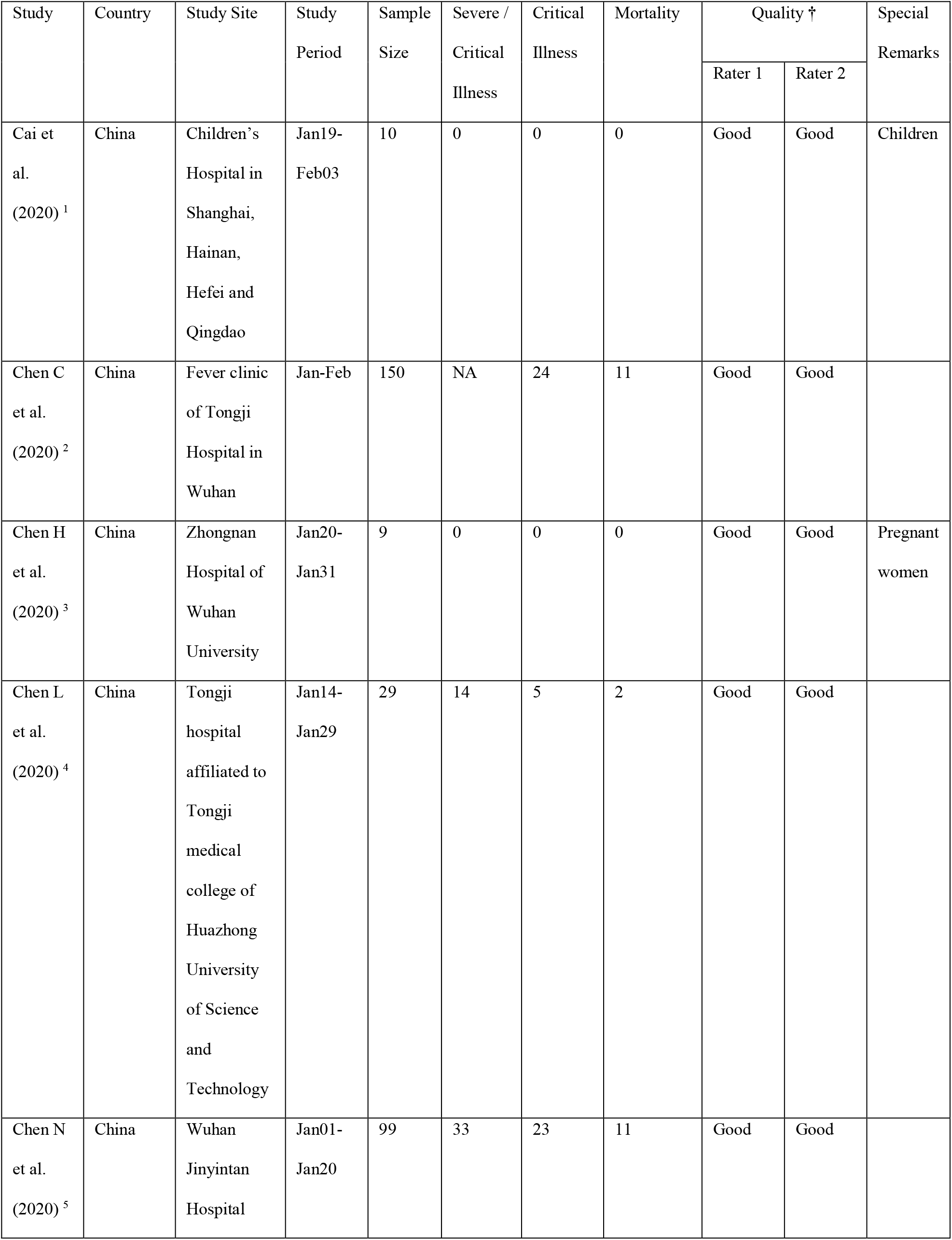
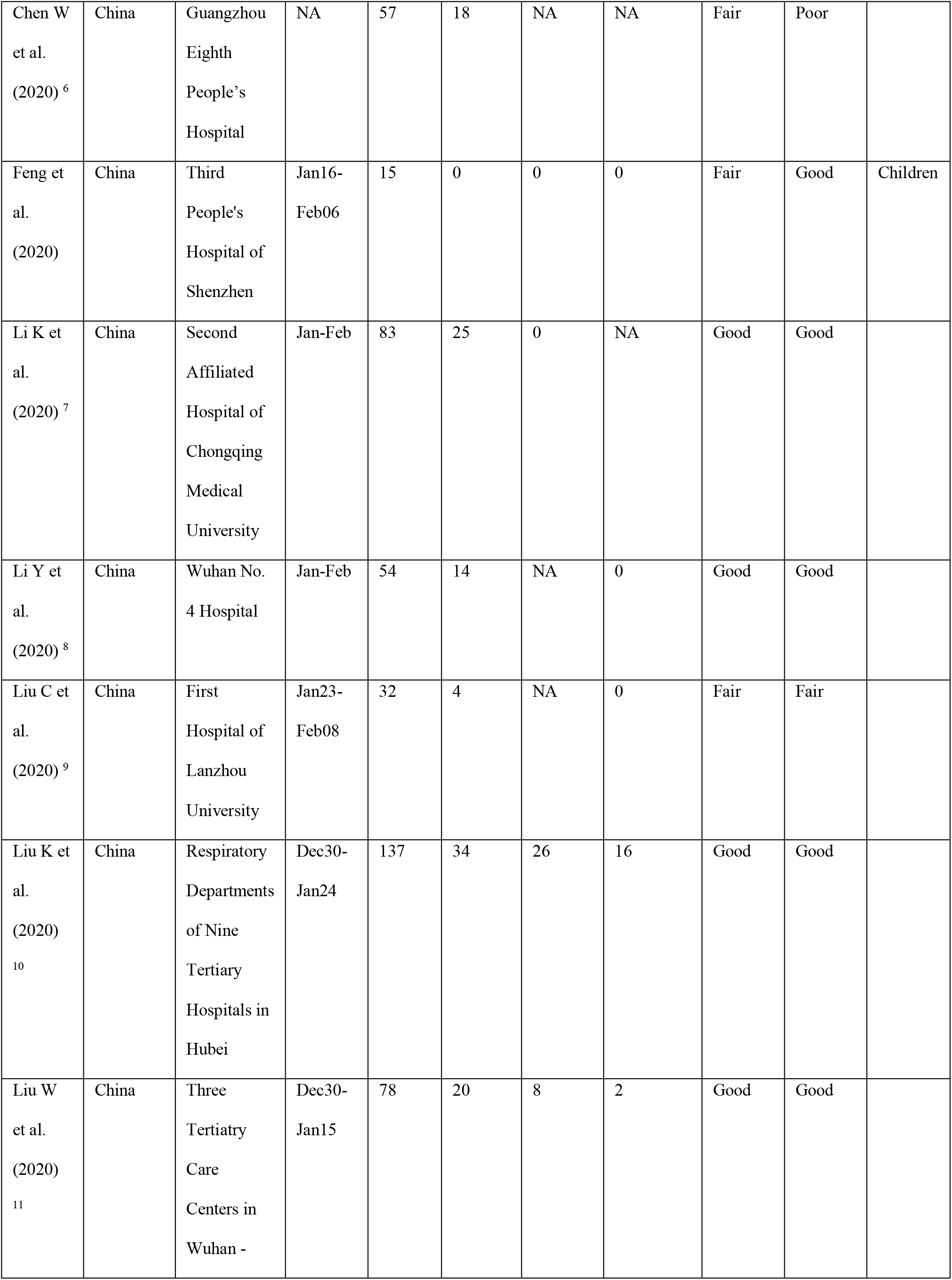
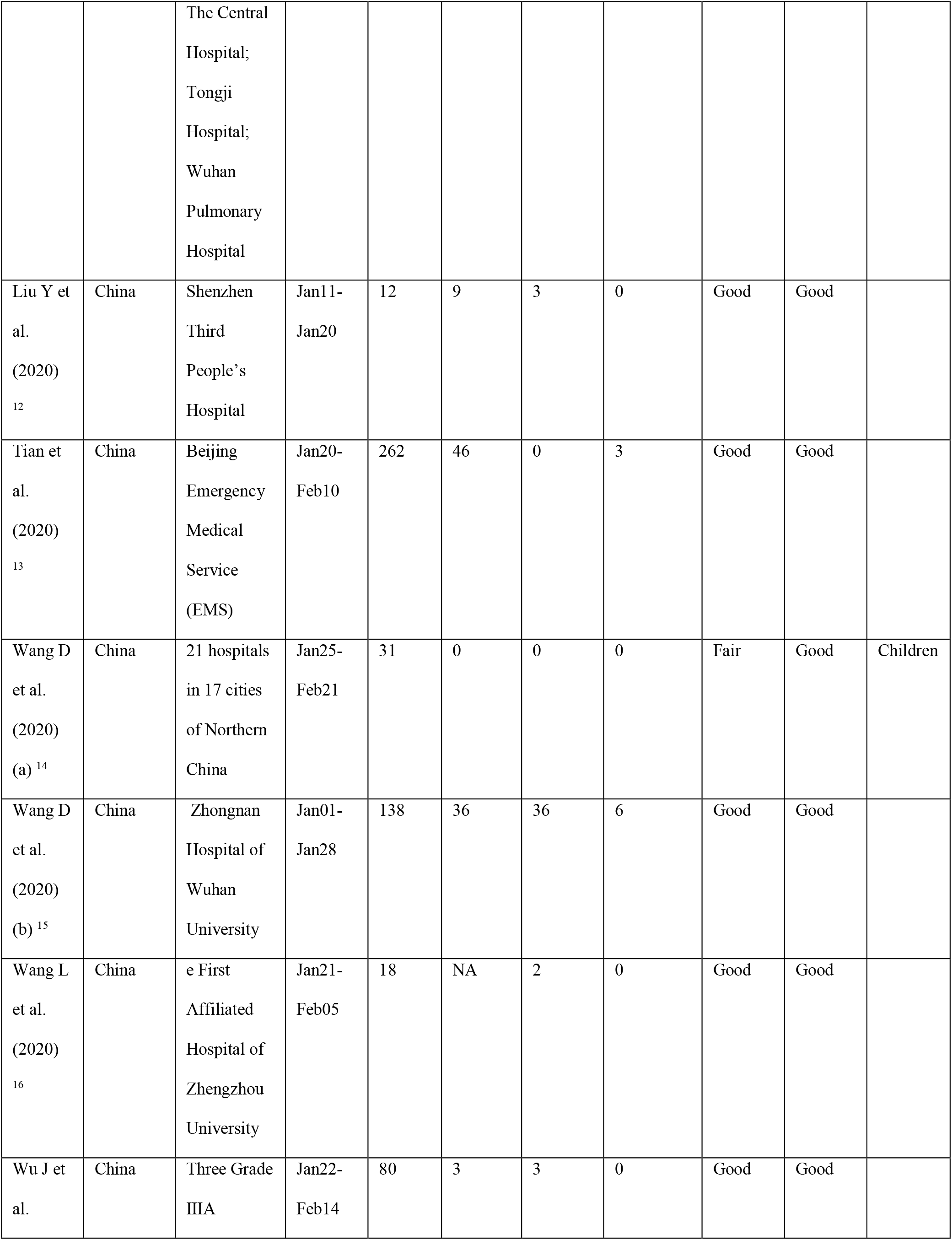
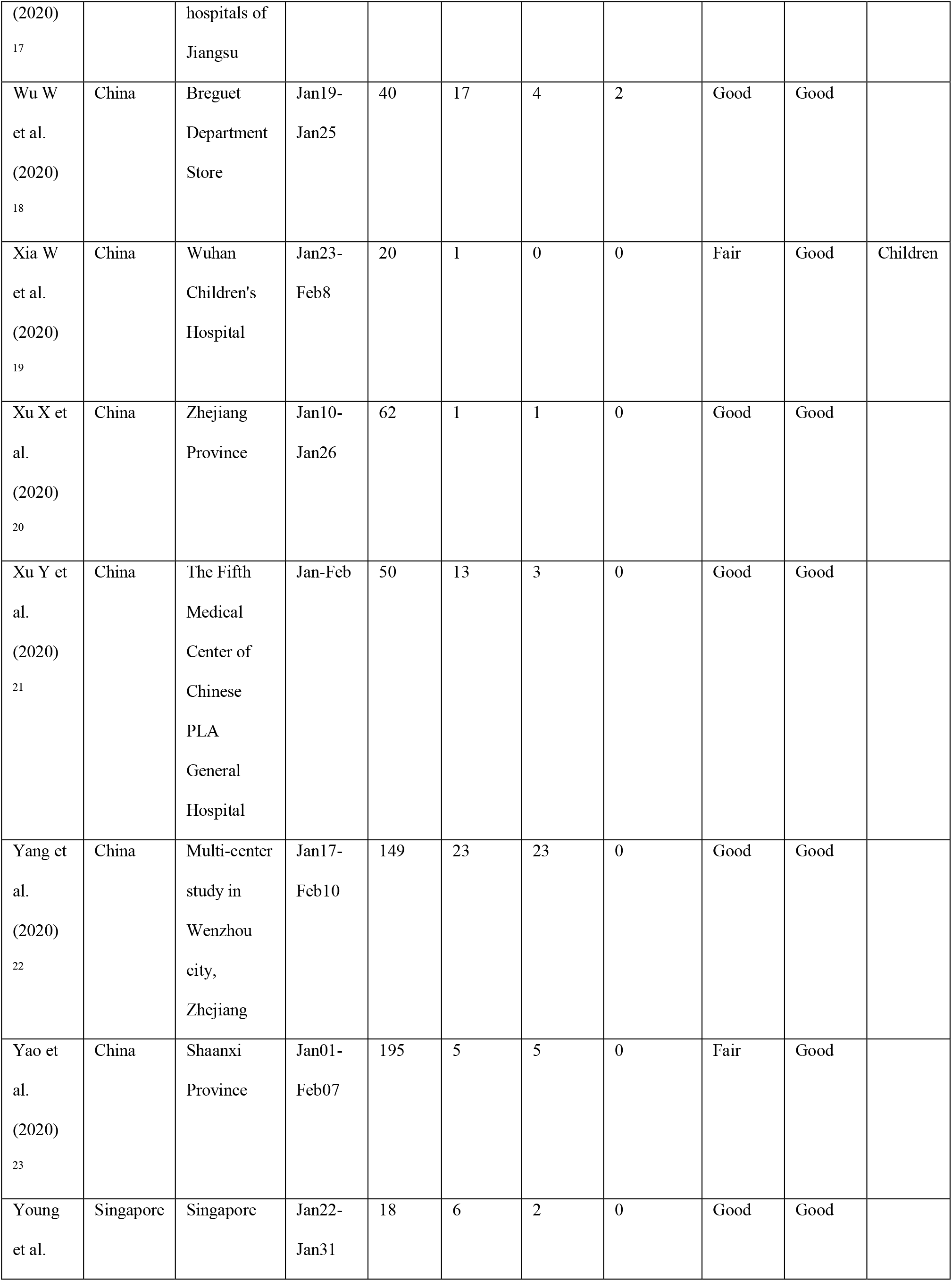
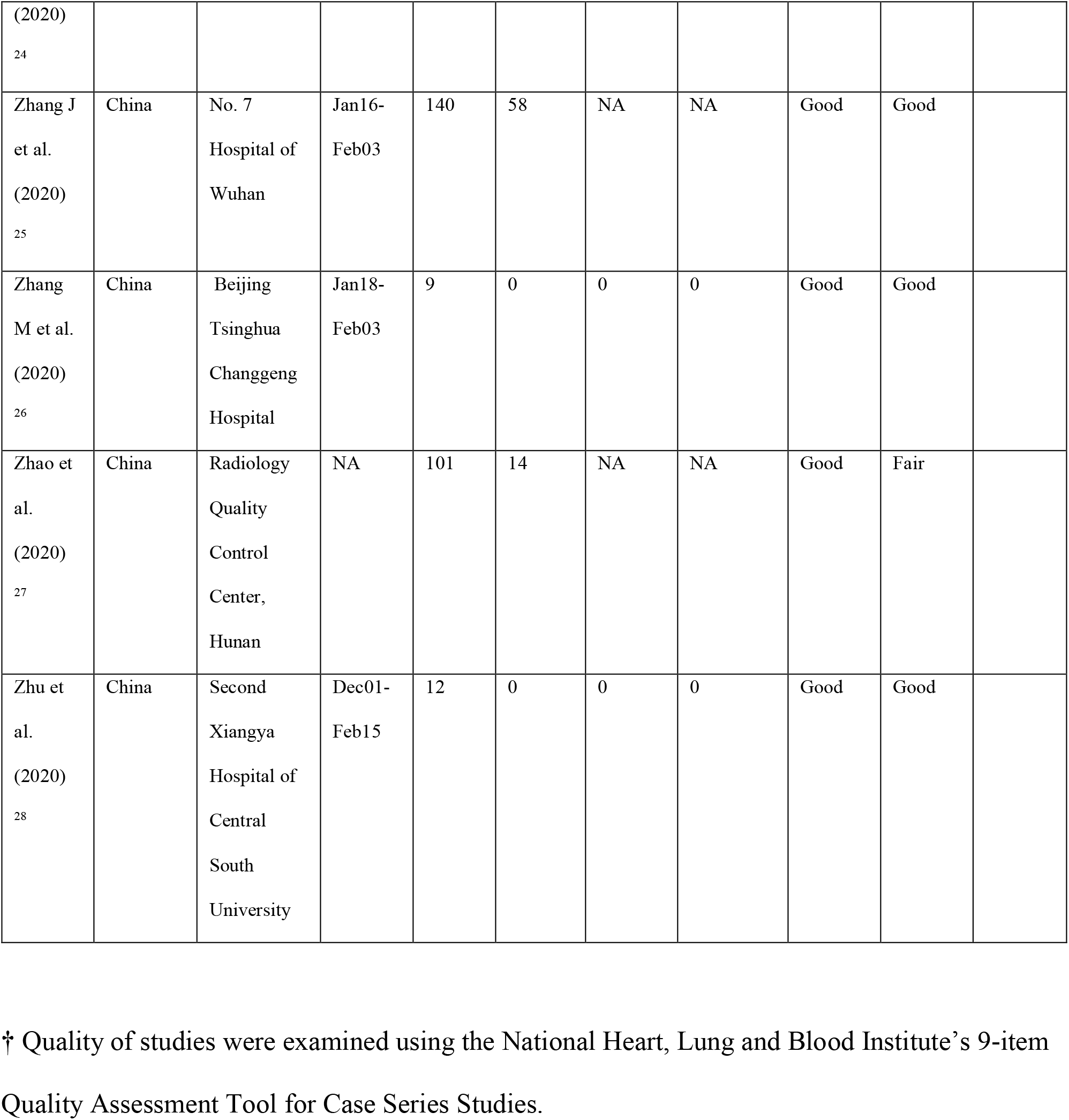
Studies meeting eligibility criteria.

**Table S3:** Summary statistics of all covariates used in the univariate meta-regression analyses.

| Variable | Events | Total | Studies | Pooled Effect Size (95%CI) † |  |
| --- | --- | --- | --- | --- | --- |
|  |  |  |  | Fixed-effects model | Random-effects model |
| Demographic variables |  |  |  |  |  |
| Age (years) | NA | 2033 | 28 | 32.847 (32.344 - 33.350) | 41.148 (32.737 - 49.558) |
| Age ≤ 18 years | 110 | 1349 | 22 | 0.082 (0.068 - 0.097) | 0.025 (0.002 - 0.274) |
| Age ≥ 60 years | 178 | 975 | 15 | 0.183 (0.160 - 0.208) | 0.064 (0.019 - 0.196) |
| Female | 941 | 2033 | 28 | 0.463 (0.441 - 0.485) | 0.466 (0.431 - 0.500) |
| Past medical history |  |  |  |  |  |
| DM-2 | 123 | 1420 | 22 | 0.087 (0.073 - 0.102) | 0.102 (0.069 - 0.148) |
| Hypertension | 228 | 1090 | 19 | 0.209 (0.186 - 0.234) | 0.120 (0.069 - 0.201) |
| Cardiac disease | 127 | 1121 | 18 | 0.113 (0.096 - 0.133) | 0.060 (0.029 - 0.120) |
| Chronic liver disease | 23 | 926 | 18 | 0.025 (0.017 - 0.037) | 0.010 (0.003 - 0.037) |
| Chronic kidney disease | 12 | 926 | 18 | 0.013 (0.007 - 0.023) | 0.013 (0.007 - 0.023) |
| Malignancy | 34 | 1216 | 19 | 0.028 (0.020 - 0.039) | 0.024 (0.014 - 0.040) |
| COPD | 19 | 1014 | 12 | 0.019 (0.012 - 0.029) | 0.018 (0.010 - 0.031) |
| Low immunity | 2 | 236 | 3 | 0.008 (0.002 - 0.033) | 0.008 (0.002 - 0.033) |
| Smoking | 16 | 247 | 3 | 0.065 (0.040 - 0.103) | 0.065 (0.040 - 0.103) |
| Pregnant | 11 | 506 | 11 | 0.022 (0.012 - 0.039) | 0.000 (0.000 - 0.386) |
| Presenting symptoms |  |  |  |  |  |
| Asymptomatic | 24 | 618 | 11 | 0.039 (0.026 - 0.057) | 0.009 (0.001 - 0.084) |
| Fever | 1310 | 1578 | 24 | 0.830 (0.811 - 0.848) | 0.833 (0.778 - 0.877) |
| Cough | 992 | 1656 | 25 | 0.599 (0.575 - 0.622) | 0.619 (0.539 - 0.693) |
| Sore throat | 121 | 909 | 16 | 0.133 (0.113 - 0.157) | 0.131 (0.085 - 0.197) |
| Tachypnea | 138 | 325 | 7 | 0.425 (0.372 - 0.479) | 0.084 (0.010 - 0.461) |
| Dyspnea | 245 | 1452 | 20 | 0.169 (0.150 - 0.189) | 0.102 (0.053 - 0.186) |
| Myalgia | 163 | 801 | 14 | 0.203 (0.177 - 0.233) | 0.192 (0.123 - 0.288) |
| Fatigue | 328 | 762 | 11 | 0.430 (0.396 - 0.466) | 0.336 (0.198 - 0.508) |
| Headache | 136 | 1253 | 16 | 0.109 (0.092 - 0.127) | 0.103 (0.071 - 0.146) |
| Diarrhea | 104 | 1288 | 21 | 0.081 (0.067 - 0.097) | 0.078 (0.059 - 0.104) |

| **Laboratory investigations ‡** |  |  |  |  |  |
| --- | --- | --- | --- | --- | --- |
| Nucleic acid test positive | 1529 | 1617 | 22 | 0.946 (0.933 - 0.956) | 1.000 (0.916 - 1.000) |
| Leukopenia | 258 | 940 | 16 | 0.274 (0.247 - 0.304) | 0.217 (0.111 - 0.380) |
| Leukocytosis | 99 | 899 | 16 | 0.110 (0.091 - 0.132) | 0.082 (0.048 - 0.137) |
| Thrombocytopenia | 73 | 472 | 8 | 0.155 (0.125 - 0.190) | 0.137 (0.078 - 0.229) |
| Lymphopenia | 476 | 988 | 17 | 0.482 (0.451 - 0.513) | 0.410 (0.289 - 0.543) |
| High LDH | 220 | 526 | 9 | 0.418 (0.377 - 0.461) | 0.433 (0.242 - 0.646) |
| Low Albumin | 156 | 423 | 6 | 0.369 (0.324 - 0.416) | 0.340 (0.063 - 0.797) |
| High CRP | 650 | 930 | 17 | 0.699 (0.669 - 0.728) | 0.668 (0.534 - 0.780) |
| High ESR | 193 | 245 | 4 | 0.788 (0.732 - 0.834) | 0.795 (0.701 - 0.866) |
| High procalcitonin | 152 | 594 | 12 | 0.256 (0.222 - 0.293) | 0.175 (0.071 - 0.370) |
| High D-dimer | 117 | 504 | 7 | 0.232 (0.197 - 0.271) | 0.188 (0.095 - 0.338) |
| **Radiological features on CT thorax** |  |  |  |  |  |
| No lesion on CT | 95 | 894 | 16 | 0.106 (0.088 - 0.128) | 0.078 (0.035 - 0.165) |
| Patchy consolidation | 231 | 551 | 11 | 0.419 (0.379 - 0.461) | 0.418 (0.314 - 0.530) |
| Ground glass opacities | 416 | 754 | 15 | 0.552 (0.516 - 0.587) | 0.693 (0.487 - 0.843) |
| Peripheral distribution | 177 | 229 | 6 | 0.773 (0.714 - 0.823) | 0.849 (0.516 - 0.967) |
| Bilateral or $\geq 3$ lobe involvement | 721 | 953 | 15 | 0.757 (0.728 - 0.783) | 0.711 (0.575 - 0.817) |

† Effect size measure is mean for age. For all other variables, it is the proportion of individuals (i.e.
events) out of total sample size.
‡ Biochemical parameters have been interpreted as low / high based on reference ranges considered in
each study.

**Table S4:** Results of all univariate meta-regression analyses examining the moderator effects of the covariates on the prevalence of combined severe or critical illness in COVID-19.

| Model | Variable | Coefficient † | SE | 95%CI | Z | p |
| --- | --- | --- | --- | --- | --- | --- |
| Demographic variables |  |  |  |  |  |  |
| 1 | Intercept | -5.921 | 1.236 | -8.344 to -3.498 | -4.789 | <0.001 |
|  | Age | 0.093 | 0.026 | 0.043 to 0.144 | 3.634 | <0.001 |
| | k = 26, $\tau^2$ = 0.781, I2 = 88.428, QM = 13.206 | | | | | |
| 2 | Intercept | -1.400 | 0.310 | -2.006 to -0.794 | -4.524 | <0.001 |
|  | Age ≤ 18 years | -3.450 | 1.287 | -5.972 to -0.927 | -2.680 | 0.007 |
| | k = 21, $\tau^2$ = 1.172, I2 = 91.709, QM = 7.182 | | | | | |
| 3 | Intercept | -3.599 | 0.461 | -4.502 to -2.695 | -7.806 | <0.001 |
|  | Age ≥ 60 years | 8.367 | 1.720 | 4.997 to 11.738 | 4.866 | <0.001 |
| | k = 15, $\tau^2$ = 0.573, I2 = 77.484, QM = 23.679 | | | | | |
| 4 | Intercept | -0.491 | 1.196 | -2.836 to 1.853 | -0.411 | 0.681 |
|  | Female | -2.836 | 2.517 | -7.769 to 2.397 | -1.127 | 0.260 |
| | k = 26, $\tau^2$ = 1.771, I2 = 94.515, QM = 1.270 | | | | | |
| Past medical history |  |  |  |  |  |  |
| 5 | Intercept | -2.344 | 0.471 | -3.267 to -1.421 | -4.978 | <0.001 |
|  | DM-2 | 3.632 | 2.161 | -0.603 to 7.868 | 1.681 | 0.093 |
| | k = 20, $\tau^2$ = 1.532, I2 = 93.418, QM = 2.826 | | | | | |
| 6 | Intercept | -2.966 | 0.556 | -4.056 to -1.877 | -5.338 | <0.001 |
|  | Hypertension | 8.381 | 2.456 | 3.567 to 13.194 | 3.413 | <0.001 |
| | k = 17, $\tau^2$ = 1.009, I2 = 88.950, QM = 11.646 | | | | | |
| 7 | Intercept | -2.112 | 0.476 | -3.045 to -1.179 | -4.436 | <0.001 |
|  | Cardiac disease | 4.801 | 2.649 | -0.392 to 9.993 | 1.812 | 0.070 |
| | k = 16, $\tau^2$ = 1.211, I2 = 91.573, QM = 3.283 | | | | | |
| 8 | Intercept | -2.274 | 0.654 | -3.555 to -0.992 | -3.476 | <0.001 |
|  | Chronic liver disease | 4.698 | 15.493 | -25.668 to 35.064 | 0.303 | 0.762 |
| | k = 16, $\tau^2$ = 3.223, I2 = 94.655, QM = 0.092 | | | | | |
| 9 | Intercept | -2.411 | 0.507 | -3.405 to -1.417 | -4.756 | <0.001 |
|  | Chronic kidney disease | 21.831 | 10.504 | 1.244 to 42.419 | 2.078 | 0.038 |
| | k = 16, $\tau^2$ = 2.235, I2 = 93.455, QM = 4.320 | | | | | |
| 10 | Intercept | -2.366 | 0.441 | -3.230 to -1.501 | -5.363 | <0.001 |
|  | Malignancy | 24.845 | 10.920 | 3.442 to 46.247 | 2.275 | 0.023 |
| | k = 17, $\tau^2$ = 0.960, I2 = 89.464, QM = 5.176 | | | | | |
| 11 | Intercept | -1.827 | 0.399 | -2.609 to -1.045 | -4.580 | <0.001 |
|  | COPD | 27.432 | 12.227 | 3.467 to 51.397 | 2.243 | 0.025 |
| | k = 11, $\tau^2$ = 0.733, I2 = 91.466, QM = 5.033 | | | | | |
| 12 | Intercept | -2.020 | 0.713 | -3.416 to -5.623 | -2.835 | 0.005 |
|  | Low immunity | 66.920 | 78.023 | -86.003 to 219.842 | 0.858 | 0.391 |
| | k = 3, $\tau^2$ = 0.733, I2 = 71.082, QM = 0.736 | | | | | |
| 13 | Intercept | -8.862 | 6.638 | -21.872 to 4.148 | -1.335 | 0.182 |
|  | Smoking | 127.878 | 101.797 | -71.641 to 327.397 | 1.256 | 0.209 |
| | k = 3, $\tau^2$ = 0.048, I2 = 50.308, QM = 1.578 | | | | | |
| 14 | Intercept | -2.088 | 0.618 | -3.299 to -2.876 | -3.377 | <0.001 |
|  | Pregnant | -4.395 | 6.052 | -16.256 to 7.466 | -0.726 | 0.468 |
| | k = 10, $\tau^2$ = 2.133, I2 = 90.869, QM = 0.527 | | | | | |
| Presenting symptoms |  |  |  |  |  |  |
| 15 | Intercept | -1.287 | 0.486 | -2.240 to -2.334 | -2.647 | 0.008 |
|  | Asymptomatic | -5.774 | 4.980 | -15.534 to 3.987 | -1.159 | 0.246 |
| | k = 11, $\tau^2$ = 1.549, I2 = 92.081, QM = 1.344 | | | | | |
| 16 | Intercept | -8.760 | 2.003 | -12.685 to -4.835 | -4.374 | <0.001 |
|  | Fever | 8.772 | 2.378 | 4.111 to 13.432 | 3.689 | <0.001 |
| | k = 23, $\tau^2$ = 0.641, I2 = 86.409, QM = 13.607 | | | | | |
| 17 | Intercept | -3.308 | 0.987 | -5.242 to -1.374 | -3.353 | <0.001 |
|  | Cough | 2.736 | 1.492 | -0.187 to 5.665 | 1.834 | 0.067 |
| | k = 24, $\tau^2$ = 1.280, I2 = 92.822, QM = 3.365 | | | | | |
| 18 | Intercept | -1.698 | 0.456 | -2.590 to -7.854 | -3.725 | <0.001 |
|  | Sore throat | -0.410 | 1.971 | -4.272 to 3.452 | -0.208 | 0.835 |
| | k = 15, $\tau^2$ = 1.035, I2 = 89.939, QM = 0.043 | | | | | |
| 19 | Intercept | -3.773 | 1.205 | -6.134 to -1.411 | -3.131 | 0.002 |
|  | Tachypnea | 5.104 | 2.616 | -0.023 to 19.231 | 1.951 | 0.051 |
| | k = 7, $\tau^2$ = 1.774, I2 = 89.572, QM = 3.806 | | | | | |
| 20 | Intercept | -1.997 | 0.354 | -2.691 to -1.322 | -5.636 | <0.001 |
|  | Dyspnea | 3.056 | 1.392 | 0.327 to 5.784 | 2.195 | 0.028 |
| | k = 19, $\tau^2$ = 0.680, I2 = 88.759, QM = 4.817 | | | | | |
| 21 | Intercept | -2.227 | 0.749 | -3.695 to -6.758 | -2.972 | 0.003 |
|  | Myalgia | 3.541 | 2.648 | -1.648 to 8.731 | 1.338 | 0.181 |
| | k = 13, $\tau^2$ = 1.323, I2 = 91.941, QM = 1.789 | | | | | |
| 22 | Intercept | -2.466 | 0.685 | -3.809 to -1.124 | -3.600 | <0.001 |
|  | Fatigue | 2.702 | 1.388 | -0.018 to 5.421 | 1.947 | 0.052 |
| | k = 11, $\tau^2$ = 0.730, I2 = 87.246, QM = 3.791 | | | | | |
| 23 | Intercept | -0.803 | 0.471 | -1.727 to 1.129 | -1.705 | 0.088 |
|  | Headache | -5.361 | 3.194 | -11.621 to 4.899 | -1.679 | 0.093 |
| | k = 15, $\tau^2$ = 1.153, I2 = 93.398, QM = 2.817 | | | | | |
| 24 | Intercept | -2.590 | 0.566 | -3.699 to -1.482 | -4.579 | <0.001 |
|  | Diarrhea | 11.680 | 5.237 | 1.415 to 21.944 | 2.230 | 0.026 |
| | k = 20, $\tau^2$ = 1.139, I2 = 91.537, QM = 4.974 | | | | | |

| Laboratory investigations ‡ |  |  |  |  |  |  |
| --- | --- | --- | --- | --- | --- | --- |
| 25 | Intercept | -0.521 | 0.404 | -1.311 to 8.272 | -1.291 | 0.197 |
|  | Nucleic acid test positive | -1.370 | 0.423 | -2.199 to -9.545 | -3.235 | 0.001 |
| | k = 21, $\tau_2$ = 1.477, I2 = 94.363, QM = 10.464 | | | | | |
| 26 | Intercept | -1.904 | 0.734 | -3.343 to -3.465 | -2.594 | 0.009 |
|  | Leukopenia | -0.357 | 1.926 | -4.132 to 3.418 | -0.185 | 0.853 |
| | k = 15, $\tau_2$ = 2.855, I2 = 96.044, QM = 0.034 | | | | | |
| 27 | Intercept | -3.166 | 0.857 | -4.845 to -1.487 | -3.696 | <0.001 |
|  | Leukocytosis | 8.608 | 5.659 | -2.482 to 19.699 | 1.521 | 0.128 |
| | k = 15, $\tau_2$ = 2.690, I2 = 95.111, QM = 2.314 | | | | | |
| 28 | Intercept | -3.945 | 1.260 | -6.414 to -1.476 | -3.132 | 0.002 |
|  | Thrombocytopenia | 10.488 | 5.507 | -0.306 to 21.282 | 1.904 | 0.057 |
| | k = 8, $\tau_2$ = 2.917, I2 = 94.454, QM = 3.627 | | | | | |
| 29 | Intercept | -3.871 | 0.840 | -5.518 to -2.224 | -4.607 | <0.001 |
|  | Lymphopenia | 4.734 | 1.574 | 1.649 to 7.823 | 3.008 | 0.003 |
| | k = 16, $\tau_2$ = 1.213, I2 = 90.850, QM = 9.046 | | | | | |
| 30 | Intercept | -4.437 | 0.686 | -5.781 to -3.192 | -6.467 | <0.001 |
|  | High LDH | 5.769 | 1.164 | 3.488 to 8.054 | 4.957 | <0.001 |
| | k = 9, $\tau_2$ = 0.278, I2 = 58.546, QM = 24.572 | | | | | |
| 31 | Intercept | -1.507 | 0.734 | -2.945 to -5.869 | -2.053 | 0.040 |
|  | Low Albumin | 1.299 | 1.384 | -1.414 to 4.312 | 0.938 | 0.348 |
| | k = 6, $\tau_2$ = 1.210, I2 = 89.774, QM = 0.880 | | | | | |
| 32 | Intercept | -4.645 | 1.018 | -6.641 to -2.649 | -4.562 | <0.001 |
|  | High CRP | 4.688 | 1.392 | 1.959 to 7.417 | 3.367 | <0.001 |
| | k = 16, $\tau_2$ = 0.697, I2 = 85.579, QM = 11.335 | | | | | |
| 33 | Intercept | 2.608 | 6.597 | -10.323 to 15.538 | 0.395 | 0.693 |
|  | High ESR | -5.846 | 8.398 | -22.305 to 11.613 | -0.696 | 0.486 |
| | k = 4, $\tau_2$ = 1.636, I2 = 84.528, QM = 0.485 | | | | | |
| 34 | Intercept | -2.535 | 0.965 | -4.427 to -0.643 | -2.626 | 0.009 |
|  | High procalcitonin | 1.146 | 2.476 | -3.707 to 5.999 | 0.463 | 0.644 |
| | k = 11, $\tau_2$ = 3.562, I2 = 95.576, QM = 0.214 | | | | | |
| 35 | Intercept | -2.962 | 0.289 | -3.528 to -2.395 | -10.253 | <0.001 |
|  | High D-dimer | 6.110 | 0.830 | 4.484 to 7.736 | 7.365 | <0.001 |
| | k = 7, $\tau_2$ = 0.000, I2 = 0.000, QM = 54.243 | | | | | |
| Radiological features on CT thorax |  |  |  |  |  |  |
| 36 | Intercept | -1.016 | 0.468 | -1.932 to -8.798 | -2.170 | 0.030 |
|  | No lesions on CT | -10.053 | 3.413 | -16.743 to -3.363 | -2.945 | 0.003 |
| | k = 15, $\tau_2$ = 1.239, I2 = 89.806, QM = 8.675 | | | | | |
| 37 | Intercept | -2.639 | 1.142 | -4.878 to -2.461 | -2.311 | 0.021 |
|  | Patchy consolidations | 1.121 | 2.265 | -3.318 to 5.559 | 0.495 | 0.621 |
| | k = 10, $\tau_2$ = 1.261, I2 = 88.147, QM = 0.245 | | | | | |
| 38 | Intercept | -2.231 | 1.134 | -4.454 to -7.938 | -1.967 | 0.049 |
|  | Ground glass opacities | 0.089 | 1.650 | -3.144 to 3.322 | 0.054 | 0.957 |
| | k = 14, $\tau^2$ = 2.583, I<sup>2</sup> = 94.256, QM = 0.003 | | | | | |
| 39 | Intercept | -6.172 | 3.565 | -13.159 to 7.816 | -1.731 | 0.083 |
|  | Peripheral distribution | 3.966 | 3.959 | -3.794 to 11.725 | 1.002 | 0.317 |
| | k = 6, $\tau^2$ = 1.767, I<sup>2</sup> = 83.815, QM = 1.003 | | | | | |
| 40 | Intercept | -4.974 | 1.267 | -7.457 to -2.492 | -3.927 | <0.001 |
| | Bilateral or ≥ 3 lobe involvement | 4.435 | 1.627 | 1.246 to 7.624 | 2.725 | 0.006 |
| | k = 14, $\tau^2$ = 0.696, I<sup>2</sup> = 87.270, QM = 7.428 | | | | | |

† Coefficients represent logit-transformed proportions; positive coefficients suggest increased risk and negative coefficients suggest decreased risk.
‡ Biochemical parameters have been interpreted as low / high based on reference ranges considered in each study.

**Table S5:** Results of all univariate meta-regression analyses examining the moderator effects of the covariates on the prevalence critical illness in COVID-19.

| Model | Variable | Coefficient † | SE | 95%CI | Z | p |
| --- | --- | --- | --- | --- | --- | --- |
| Demographic variables |  |  |  |  |  |  |
| 1 | Intercept | -8.153 | 2.031 | -12.134 to -4.173 | -4.014 | <0.001 |
|  | Age | 0.115 | 0.041 | 0.033 to 6.196 | 2.774 | 0.006 |
| | k = 24, $\tau^2$ = 1.094, I2 = 85.333, QM = 7.694 | | | | | |
| 2 | Intercept | -2.407 | 0.527 | -3.440 to -1.374 | -4.567 | <0.001 |
| | Age ≤ 18 years | -7.526 | 6.892 | -21.034 to 5.981 | -1.092 | 0.275 |
| | k = 18, $\tau^2$ = 1.635, I2 = 87.100, QM = 1.193 | | | | | |
| 3 | Intercept | -5.479 | 0.884 | -7.211 to -3.747 | -6.200 | <0.001 |
| | Age ≥ 60 years | 8.868 | 2.647 | 3.678 to 14.757 | 3.349 | <0.001 |
| | k = 13, $\tau^2$ = 0.981, I2 = 69.156, QM = 11.219 | | | | | |
| 4 | Intercept | -1.157 | 1.429 | -3.957 to 1.644 | -0.809 | 0.418 |
|  | Female | -3.934 | 3.082 | -9.975 to 2.157 | -1.276 | 0.202 |
| | k = 24, $\tau^2$ = 1.890, I2 = 90.774, QM = 1.629 | | | | | |
| Past medical history |  |  |  |  |  |  |
| 5 | Intercept | -2.712 | 0.510 | -3.711 to -1.713 | -5.319 | <0.001 |
|  | DM-2 | 0.581 | 2.220 | -3.770 to 4.932 | 0.262 | 0.794 |
| | k = 19, $\tau^2$ = 1.405, I2 = 89.782, QM = 0.068 | | | | | |
| 6 | Intercept | -4.091 | 0.676 | -5.416 to -2.765 | -6.050 | <0.001 |
|  | Hypertension | 8.385 | 2.518 | 3.450 to 13.321 | 3.330 | <0.001 |
| | k = 17, $\tau^2$ = 0.615, I2 = 74.934, QM = 11.090 | | | | | |
| 7 | Intercept | -3.386 | 0.657 | -4.674 to -2.597 | -5.150 | <0.001 |
|  | Cardiac disease | 6.745 | 2.970 | 0.924 to 12.565 | 2.271 | 0.023 |
| | k = 15, $\tau^2$ = 1.058, I2 = 86.342, QM = 5.158 | | | | | |
| 8 | Intercept | -2.632 | 0.494 | -3.600 to -1.664 | -5.329 | <0.001 |
|  | Chronic liver disease | -1.965 | 11.525 | -24.553 to 21.623 | -0.170 | 0.865 |
| | k = 15, $\tau^2$ = 1.332, I2 = 84.529, QM = 0.029 | | | | | |
| 9 | Intercept | -2.850 | 0.463 | -3.757 to -1.944 | -6.161 | <0.001 |
|  | Chronic kidney disease | 11.613 | 8.335 | -4.722 to 27.949 | 1.393 | 0.163 |
| | k = 15, $\tau^2$ = 1.185, I2 = 84.105, QM = 1.942 | | | | | |
| 10 | Intercept | -2.718 | 0.428 | -3.556 to -1.886 | -6.355 | <0.001 |
|  | Malignancy | 20.308 | 9.919 | 0.866 to 39.752 | 2.047 | 0.041 |
| | k = 16, $\tau^2$ = 0.497, I2 = 75.460, QM = 4.191 | | | | | |
| 11 | Intercept | -2.140 | 0.468 | -3.057 to -1.223 | -4.573 | <0.001 |
|  | COPD | -3.422 | 14.786 | -32.402 to 25.557 | -0.231 | 0.817 |
| | k = 11, $\tau^2$ = 1.028, I2 = 89.416, QM = 0.054 | | | | | |

| Presenting symptoms |  |  |  |  |  |  |
| --- | --- | --- | --- | --- | --- | --- |
| 12 | Intercept | -3.186 | 1.049 | -5.243 to -1.129 | -3.036 | 0.002 |
|  | Asymptomatic | -9.113 | 12.896 | -34.389 to 16.162 | -0.707 | 0.480 |
| | k = 9, $\tau_2$ = 4.359, I2 = 90.352, QM = 0.499 | | | | | |
| 13 | Intercept | -10.098 | 3.624 | -17.201 to -2.995 | -2.786 | 0.005 |
|  | Fever | 8.525 | 4.236 | 0.224 to 16.827 | 2.013 | 0.044 |
| | k = 20, $\tau_2$ = 1.946, I2 = 89.018, QM = 4.051 | | | | | |
| 14 | Intercept | -4.429 | 1.361 | -7.096 to -1.762 | -3.255 | 0.001 |
|  | Cough | 2.421 | 2.119 | -1.732 to 6.574 | 1.142 | 0.253 |
| | k = 21, $\tau_2$ = 2.096, I2 = 90.704, QM = 1.305 | | | | | |
| 15 | Intercept | -2.901 | 0.617 | -4.110 to -1.693 | -4.706 | <0.001 |
|  | Sore throat | -0.011 | 2.573 | -5.054 to 5.733 | -0.004 | 0.997 |
| | k = 14, $\tau_2$ = 1.552, I2 = 87.761, QM = 0.000 | | | | | |
| 16 | Intercept | -6.493 | 3.574 | -13.498 to 6.513 | -1.816 | 0.069 |
|  | Tachypnea | 4.139 | 6.653 | -8.901 to 17.179 | 0.622 | 0.534 |
| | k = 7, $\tau_2$ = 9.745, I2 = 92.103, QM = 0.387 | | | | | |
| 17 | Intercept | -3.919 | 0.701 | -5.293 to -2.546 | -5.592 | <0.001 |
|  | Dyspnea | 5.263 | 2.589 | 0.187 to 19.339 | 2.033 | 0.042 |
| | k = 17, $\tau_2$ = 1.935, I2 = 90.510, QM = 4.131 | | | | | |
| 18 | Intercept | -2.954 | 0.729 | -4.382 to -1.526 | -4.055 | <0.001 |
|  | Myalgia | 2.311 | 2.833 | -3.241 to 7.864 | 0.816 | 0.415 |
| | k = 13, $\tau_2$ = 1.128, I2 = 82.759, QM = 0.666 | | | | | |
| 19 | Intercept | -5.730 | 1.452 | -8.575 to -2.884 | -3.947 | <0.001 |
|  | Fatigue | 6.841 | 3.255 | 0.462 to 13.221 | 2.102 | 0.036 |
| | k = 9, $\tau_2$ = 1.883, I2 = 75.036, QM = 4.418 | | | | | |
| 20 | Intercept | -2.093 | 0.707 | -3.478 to -4.718 | -2.962 | 0.003 |
|  | Headache | -7.333 | 6.010 | -19.113 to 4.446 | -1.220 | 0.222 |
| | k = 15, $\tau_2$ = 1.972, I2 = 92.662, QM = 1.489 | | | | | |
| 21 | Intercept | -3.018 | 0.673 | -4.337 to -1.699 | -4.485 | <0.001 |
|  | Diarrhea | 4.228 | 5.997 | -7.525 to 15.981 | 0.705 | 0.481 |
| | k = 18, $\tau_2$ = 1.279, I2 = 86.591, QM = 0.497 | | | | | |
| Laboratory investigations ‡ |  |  |  |  |  |  |
| 22 | Intercept | -728.908 | 26694.139 | -53048.459 to 51594.644 | -0.027 | 0.978 |
|  | Nucleic acid test positive | 725.702 | 26694.139 | -51593.849 to 53945.253 | 0.027 | 0.978 |
| | k = 18, $\tau_2$ = 2.861, I2 = 94.023, QM = 0.001 | | | | | |
| 23 | Intercept | -2.923 | 0.688 | -4.272 to -1.575 | -4.249 | <0.001 |
|  | Leukopenia | -0.149 | 1.679 | -3.440 to 3.141 | -0.089 | 0.929 |
| | k = 15, $\tau_2$ = 1.763, I2 = 87.768, QM = 0.008 | | | | | |
| 24 | Intercept | -3.734 | 0.742 | -5.189 to -2.279 | -5.030 | <0.001 |
|  | Leukocytosis | 6.462 | 4.651 | -2.655 to 15.578 | 1.389 | 0.165 |
| | k = 15, $\tau_2$ = 1.617, I2 = 83.727, QM = 1.930 | | | | | |
| 25 | Intercept | -3.319 | 0.906 | -5.095 to -1.543 | -3.663 | <0.001 |
|  | Thrombocytopenia | 4.623 | 3.915 | -3.049 to 12.296 | 1.181 | 0.238 |
| | k = 8, $\tau_2$ = 1.198, I2 = 85.576, QM = 1.395 | | | | | |
| 26 | Intercept | -4.303 | 0.979 | -6.222 to -2.383 | -4.393 | <0.001 |
|  | Lymphopenia | 3.533 | 1.996 | -0.378 to 7.444 | 1.770 | 0.077 |
| | k = 15, $\tau_2$ = 1.264, I2 = 81.627, QM = 3.134 | | | | | |
| 27 | Intercept | -3.882 | 0.707 | -5.267 to -2.496 | -5.491 | <0.001 |
|  | High LDH | 3.392 | 1.137 | 1.164 to 5.621 | 2.984 | 0.003 |
| | k = 8, $\tau_2$ = 0.315, I2 = 51.385, QM = 8.904 | | | | | |
| 28 | Intercept | -2.131 | 0.308 | -2.735 to -1.527 | -6.914 | <0.001 |
|  | Low Albumin | 0.994 | 0.517 | -0.019 to 2.318 | 1.923 | 0.055 |
| | k = 5, $\tau_2$ = 0.081, I2 = 27.279, QM = 3.697 | | | | | |
| 29 | Intercept | -6.561 | 1.654 | -9.803 to -3.319 | -3.967 | <0.001 |
|  | High CRP | 5.696 | 2.155 | 1.471 to 9.921 | 2.642 | 0.008 |
| | k = 14, $\tau_2$ = 0.741, I2 = 74.102, QM = 6.983 | | | | | |
| 30 | Intercept | -2.584 | 0.805 | -4.161 to -1.936 | -3.209 | 0.001 |
|  | High procalcitonin | -6.018 | 4.132 | -14.116 to 2.081 | -1.456 | 0.145 |
| | k = 10, $\tau_2$ = 2.030, I2 = 78.770, QM = 2.121 | | | | | |
| 31 | Intercept | -3.367 | 0.736 | -4.808 to -1.925 | -4.577 | <0.001 |
|  | High D-dimer | 6.185 | 2.671 | 0.950 to 11.421 | 2.315 | 0.021 |
| | k = 5, $\tau_2$ = 0.221, I2 = 39.400, QM = 5.361 | | | | | |
| Radiological features on CT thorax |  |  |  |  |  |  |
| 32 | Intercept | -2.481 | 0.621 | -3.698 to -1.265 | -3.998 | <0.001 |
|  | No lesions on CT | -6.518 | 3.726 | -13.821 to 2.786 | -1.749 | 0.080 |
| | k = 14, $\tau_2$ = 1.473, I2 = 79.098, QM = 3.059 | | | | | |
| 33 | Intercept | -2.430 | 1.574 | -5.514 to 9.656 | -1.544 | 0.123 |
|  | Patchy consolidations | -2.949 | 3.633 | -10.070 to 4.171 | -0.812 | 0.417 |
| | k = 10, $\tau_2$ = 2.026, I2 = 77.274, QM = 0.659 | | | | | |
| 34 | Intercept | -1.955 | 0.893 | -3.705 to -6.204 | -2.188 | 0.029 |
|  | Ground glass opacities | -1.912 | 1.387 | -4.630 to 8.856 | -1.379 | 0.168 |
| | k = 14, $\tau_2$ = 1.354, I2 = 81.205, QM = 1.901 | | | | | |
| 35 | Intercept | -5.393 | 3.288 | -11.838 to 1.452 | -1.640 | 0.101 |
|  | Peripheral distribution | 1.619 | 3.529 | -5.297 to 8.536 | 0.459 | 0.646 |
| | k = 5, $\tau_2$ = 0.617, I2 = 32.942, QM = 0.211 | | | | | |
| 36 | Intercept | -4.371 | 1.735 | -7.772 to -1.978 | -2.519 | 0.012 |
| | Bilateral or $\geq 3$ lobe involvement | 1.857 | 2.445 | -2.935 to 6.657 | 0.759 | 0.448 |
| | k = 12, $\tau_2$ = 1.547, I2 = 84.184, QM = 0.577 | | | | | |

† Coefficients represent logit-transformed proportions; positive coefficients suggest increased risk and negative coefficients suggest decreased risk.
‡ Biochemical parameters have been interpreted as low / high based on reference ranges considered in each study.

**Table S6:** Results of all univariate meta-regression analyses examining the moderator effects of the covariates on the case-fatality rate in COVID-19.

| Model | Variable | Coefficient † | SE | 95%CI | Z | p |
| --- | --- | --- | --- | --- | --- | --- |
| Demographic variables |  |  |  |  |  |  |
| 1 | Intercept | -13.847 | 2.681 | -19.101 to -8.593 | -5.165 | <0.001 |
|  | Age | 0.198 | 0.052 | 0.096 to 9.299 | 3.838 | <0.001 |
| | k = 25, $\tau_2 = 0.506$, I2 = 50.639, QM = 14.733 | | | | | |
| 2 | Intercept | -3.250 | 0.553 | -4.334 to -2.166 | -5.874 | <0.001 |
|  | Age ≤ 18 years | -39.307 | 18.722 | -76.002 to -2.613 | -2.100 | 0.036 |
| | k = 20, $\tau_2 = 0.755$, I2 = 57.711, QM = 4.408 | | | | | |
| 3 | Intercept | -6.156 | 1.012 | -8.140 to -4.172 | -6.081 | <0.001 |
|  | Age ≥ 60 years | 7.499 | 2.949 | 1.720 to 13.278 | 2.543 | 0.011 |
| | k = 14, $\tau_2 = 0.731$, I2 = 44.403, QM = 6.468 | | | | | |
| 4 | Intercept | -4.422 | 1.938 | -8.221 to -3.623 | -2.281 | 0.023 |
|  | Female | -0.868 | 3.930 | -8.570 to 6.834 | -0.221 | 0.825 |
| | k = 25, $\tau_2 = 2.986$, I2 = 84.794, QM = 0.049 | | | | | |
| Past medical history |  |  |  |  |  |  |
| 5 | Intercept | -3.966 | 0.865 | -5.660 to -2.271 | -4.587 | <0.001 |
|  | DM-2 | -4.139 | 4.769 | -13.485 to 5.257 | -0.868 | 0.385 |
| | k = 19, $\tau_2 = 2.493$, I2 = 85.131, QM = 0.754 | | | | | |
| 6 | Intercept | -4.843 | 0.937 | -6.680 to -3.396 | -5.167 | <0.001 |
|  | Hypertension | 6.070 | 3.098 | -0.002 to 12.143 | 1.959 | 0.050 |
| | k = 17, $\tau_2 = 0.818$, I2 = 62.909, QM = 3.839 | | | | | |
| 7 | Intercept | -5.658 | 1.528 | -8.653 to -2.663 | -3.703 | <0.001 |
|  | Cardiac disease | 6.298 | 5.635 | -4.746 to 17.343 | 1.118 | 0.264 |
| | k = 16, $\tau_2 = 3.590$, I2 = 87.325, QM = 1.249 | | | | | |
| 8 | Intercept | -5.096 | 1.329 | -7.701 to -2.491 | -3.833 | <0.001 |
|  | Chronic liver disease | 2.410 | 21.686 | -40.093 to 44.913 | 0.111 | 0.912 |
| | k = 17, $\tau_2 = 3.515$, I2 = 84.824, QM = 0.012 | | | | | |
| 9 | Intercept | -4.976 | 1.169 | -7.267 to -2.685 | -4.256 | <0.001 |
|  | Chronic kidney disease | -4.095 | 24.503 | -52.120 to 43.938 | -0.167 | 0.867 |
| | k = 17, $\tau_2 = 3.415$, I2 = 85.031, QM = 0.028 | | | | | |
| 10 | Intercept | -5.202 | 1.169 | -7.492 to -2.912 | -4.452 | <0.001 |
|  | Malignancy | 25.193 | 22.794 | -19.482 to 69.869 | 1.105 | 0.269 |
| | k = 18, $\tau_2 = 2.874$, I2 = 86.440, QM = 1.222 | | | | | |
| 11 | Intercept | -4.718 | 1.206 | -7.081 to -2.355 | -3.913 | <0.001 |
|  | COPD | 11.975 | 37.673 | -61.863 to 85.813 | 0.318 | 0.751 |
| | k = 10, $\tau_2 = 3.332$, I2 = 90.202, QM = 0.101 | | | | | |

| Presenting symptoms |  |  |  |  |  |  |
| --- | --- | --- | --- | --- | --- | --- |
| 12 | Intercept | -3.582 | 0.815 | -5.179 to -1.985 | -4.397 | <0.001 |
|  | Asymptomatic | -22.825 | 23.869 | -69.607 to 23.957 | -0.956 | 0.339 |
| | k = 10, $\tau^2$ = 0.832, I2 = 53.265, QM = 0.914 | | | | | |
| 13 | Intercept | -16.665 | 6.768 | -29.930 to -3.401 | -2.462 | 0.014 |
|  | Fever | 13.979 | 7.552 | -0.823 to 28.781 | 1.851 | 0.064 |
| | k = 21, $\tau^2$ = 2.575, I2 = 81.750, QM = 3.426 | | | | | |
| 14 | Intercept | -4.380 | 1.630 | -7.574 to -1.186 | -2.688 | 0.007 |
|  | Cough | -0.590 | 2.693 | -5.867 to 4.688 | -0.219 | 0.827 |
| | k = 22, $\tau^2$ = 2.526, I2 = 79.906, QM = 0.048 | | | | | |
| 15 | Intercept | -4.031 | 1.764 | -7.488 to -9.574 | -2.285 | 0.022 |
|  | Sore throat | -12.722 | 13.745 | -39.662 to 14.219 | -0.926 | 0.355 |
| | k = 14, $\tau^2$ = 3.976, I2 = 82.831, QM = 0.857 | | | | | |
| 16 | Intercept | -4.512 | 1.018 | -6.507 to -2.518 | -4.434 | <0.001 |
|  | Tachypnea | 3.019 | 2.240 | -1.371 to 7.419 | 1.348 | 0.178 |
| | k = 6, $\tau^2$ = 0.000, I2 = 0.000, QM = 1.816 | | | | | |
| 17 | Intercept | -5.384 | 0.963 | -7.272 to -3.497 | -5.592 | <0.001 |
|  | Dyspnea | 5.208 | 2.959 | -0.592 to 11.528 | 1.760 | 0.078 |
| | k = 17, $\tau^2$ = 1.833, I2 = 78.737, QM = 3.097 | | | | | |
| 18 | Intercept | -5.809 | 1.825 | -9.386 to -2.232 | -3.183 | 0.001 |
|  | Myalgia | 3.859 | 5.323 | -6.575 to 14.292 | 0.725 | 0.469 |
| | k = 13, $\tau^2$ = 3.173, I2 = 78.683, QM = 0.525 | | | | | |
| 19 | Intercept | -4.991 | 0.697 | -6.357 to -3.625 | -7.162 | <0.001 |
|  | Fatigue | 2.601 | 1.314 | 0.025 to 5.178 | 1.979 | 0.048 |
| | k = 10, $\tau^2$ = 0.000, I2 = 0.000, QM = 3.917 | | | | | |
| 20 | Intercept | -3.100 | 0.975 | -5.010 to -1.189 | -3.180 | 0.001 |
|  | Headache | -13.603 | 10.138 | -33.474 to 6.268 | -1.342 | 0.180 |
| | k = 15, $\tau^2$ = 2.078, I2 = 83.381, QM = 1.800 | | | | | |
| 21 | Intercept | -5.158 | 1.600 | -8.294 to -2.223 | -3.224 | 0.001 |
|  | Diarrhea | 0.674 | 12.728 | -24.272 to 25.629 | 0.053 | 0.958 |
| | k = 18, $\tau^2$ = 4.168, I2 = 87.268, QM = 0.003 | | | | | |
| Laboratory investigations ‡ |  |  |  |  |  |  |
| 22 | Intercept | -277.080 | 203424.495 | -398981.762 to 398427.693 | -0.001 | 0.999 |
|  | Nucleic acid test positive | 272.246 | 203424.495 | -398432.438 to 398976.929 | 0.001 | 0.999 |
| | k = 18, $\tau^2$ = 2.967, I2 = 85.532, QM = 0.000 | | | | | |
| 23 | Intercept | -5.304 | 2.008 | -9.239 to -1.369 | -2.642 | 0.008 |
|  | Leukopenia | -2.851 | 4.864 | -12.385 to 6.682 | -0.586 | 0.558 |
| | k = 14, $\tau^2$ = 7.059, I2 = 89.937, QM = 0.344 | | | | | |
| 24 | Intercept | -7.326 | 1.868 | -10.987 to -3.665 | -3.922 | <0.001 |
|  | Leukocytosis | 20.497 | 7.588 | 5.624 to 35.369 | 2.701 | 0.007 |
| | k = 14, $\tau^2$ = 0.790, I2 = 55.410, QM = 7.296 | | | | | |
| 25 | Intercept | -5.628 | 2.680 | -10.880 to -3.375 | -2.100 | 0.036 |
|  | Thrombocytopenia | -2.645 | 12.249 | -26.652 to 21.362 | -0.216 | 0.829 |
| | k = 8, $\tau^2$ = 7.292, I2 = 89.313, QM = 0.047 | | | | | |
| 26 | Intercept | -10.089 | 4.017 | -17.961 to -2.216 | -2.512 | 0.012 |
|  | Lymphopenia | 8.553 | 6.098 | -3.398 to 24.574 | 1.403 | 0.161 |
| | k = 15, $\tau^2$ = 6.297, I2 = 87.338, QM = 1.967 | | | | | |
| 27 | Intercept | -9.907 | 3.336 | -16.444 to -3.369 | -2.970 | 0.003 |
|  | High LDH | 8.923 | 4.123 | 0.842 to 17.604 | 2.164 | 0.030 |
| | k = 9, $\tau^2$ = 0.938, I2 = 46.607, QM = 4.684 | | | | | |
| 28 | Intercept | -6.740 | 1.325 | -9.336 to -4.143 | -5.088 | <0.001 |
|  | Low Albumin | 4.782 | 1.433 | 1.973 to 7.591 | 3.337 | <0.001 |
| | k = 6, $\tau^2$ = 0.000, I2 = 0.000, QM = 11.136 | | | | | |
| 29 | Intercept | -22.970 | 10.385 | -43.324 to -2.616 | -2.212 | 0.027 |
|  | High CRP | 23.196 | 11.885 | -0.097 to 46.499 | 1.952 | 0.051 |
| | k = 15, $\tau^2$ = 1.120, I2 = 63.919, QM = 3.809 | | | | | |
| 30 | Intercept | -6.882 | 4.270 | -15.250 to 1.486 | -1.612 | 0.107 |
|  | High procalcitonin | -12.881 | 22.821 | -57.609 to 31.847 | -0.564 | 0.572 |
| | k = 10, $\tau^2$ = 11.734, I2 = 89.835, QM = 0.319 | | | | | |
| Radiological features on CT thorax |  |  |  |  |  |  |
| 32 | Intercept | -2.321 | 0.316 | -2.939 to -1.791 | -7.344 | <0.001 |
|  | No lesions on CT | -1941.342 | 3948.493 | -9680.245 to 5797.561 | -0.492 | 0.623 |
| | k = 13, $\tau^2$ = 0.000, I2 = 0.000, QM = 0.242 | | | | | |
| 33 | Intercept | -4.454 | 2.631 | -9.609 to 2.702 | -1.693 | 0.090 |
|  | Patchy consolidations | -1.290 | 5.246 | -11.572 to 8.992 | -0.246 | 0.806 |
| | k = 9, $\tau^2$ = 2.859, I2 = 72.114, QM = 0.060 | | | | | |
| 34 | Intercept | -1.341 | 1.554 | -4.387 to 1.746 | -0.863 | 0.388 |
|  | Ground glass opacities | -8.374 | 5.039 | -18.250 to 1.552 | -1.662 | 0.097 |
| | k = 13, $\tau^2$ = 2.225, I2 = 77.488, QM = 2.762 | | | | | |
| 35 | Intercept | -9.953 | 4.723 | -19.210 to -2.695 | -2.107 | 0.035 |
| | Bilateral or $\geq 3$ lobe involvement | 7.011 | 6.069 | -4.883 to 18.956 | 1.155 | 0.248 |
| | k = 12, $\tau^2$ = 3.840, I2 = 86.622, QM = 1.335 | | | | | |

† Coefficients represent logit-transformed proportions; positive coefficients suggest increased risk and negative coefficients suggest decreased risk.
‡ Biochemical parameters have been interpreted as low / high based on reference ranges considered in each study.

